# Geometry based gene expression signatures detect cancer treatment responders in clinical trials

**DOI:** 10.1101/2024.07.01.24309803

**Authors:** Wojciech Chachólski, Ryan Ramanujam

## Abstract

**Aim:** The overall aim of this project is to determine if gene expression signatures of tumors, constructed from geometrical attributes of data, can be used to both create a definitive classification of responders and non-responders, and to predict patient treatment response in an unbiased manner. This is tested in an open-sourced Pfizer clinical trial data on avelumab plus axitnib in advanced renal cell carcinoma (*n* = 726).

**Results:** Geometrical gene expression signatures were able to be used to create standardized classification of responders to the intervention, as demonstrated by dramatically different Kaplan-Meier (KM) estimators based on responder category assigned in the Pfizer trial. Furthermore, unbiased prediction based on leave one out methodology was able to correctly predict the responder classification with 82.0% accuracy. Biomarkers of response generated indicated that the strongest predictive gene was PODXL (podocalyxin), with an inconsistent influence on responder class based on over- and underexpression. A KM estimator of the out-of-sample predictions showed nearly four times the average effect in samples predicted to be responders against those predicted to not be responders, and accounted for 79.2% of the treatment effect.

**Conclusions:** Gene expression based geometrical signatures are able to create “gold standard” classification of responders and non-responders in clinical trial data, and are highly accurate at predicting these labels in an out-of-sample, unbiased test.

These methods can be used to find more stable biomarkers of response, as well as increase the chances of a clinical trial being approved.

## 1 Introduction

Precision medicine has the goal of individualizing the treatment for a patient based on that patient’s particular characteristics, which may include data on patient demographics and clinical information, as well as experimental data such as biomarkers, genomics, gene expression, proteomics, metabolomics, etc. By using individual response to treatment, a more efficacious treatment approach or compound can be selected based on analysis of this data [1]. In oncology, tumors can be characterized by molecular approaches such as measuring gene expression or mutations. The outcomes for each patient in clinical studies are typically reflected in time to event analysis, with increased time to progression-free survival (PFS) being frequently used to indicate better treatment effect.

Mutations are currently the most widely used biomarker for dividing patients and tumors into clinical subgroups [2]. This may be a single somatic mutation or a combination of several mutations. This has led to efforts to find biomarkers across tumor types which may indicate improved treatment response [3]. The ability to detect individuals who may respond to a treatment across various tumor tissues would have strong clinical utility for precision oncology [4]. Nevertheless, identifying such biomarkers to detect a large proportion of patients with efficacious responses remains difficult [1].

Some cancer therapies have been FDA approved based on the presence of certain mutations [5]. However, data from the Dependency Map project (DepMap) [6] presented evidence that gene expression is the data type most strongly associated with treatment response. Given this, efforts to combine gene expression data into signatures which correspond to tumor biology and/or state are warranted. One method of combining complex data, such as from RNA-seq experiments, is by using techniques being developed in geometrical and topological analysis.

Topological data analysis (TDA) is a rapidly growing field enabling the ability to structure and organize data based on its geometry. It transforms data into invariants that capture higher order interactions and are amenable for statistical and machine learning analysis. The presence of the higher order information often can yield insights which are not available with standard statistical methods. This field has been applied to biomedical research, with results in breast cancer characterizing tumors with unique characteristics [7]. It also was used to identify Type 2 diabetes (T2D) subgroups based on clinical features which had distinct genetic associations [8].

We have developed several TDA methods based on classical TDA approaches such as extending the mapper algorithm for multi-measurements [9], and creating a new invariant called stable rank [10, 11] which is a key component of the technology which is presented here. Stable rank has also been used to create supervised learning methods [12], which have been then applied to characterization of sex-based differences in microglia [13]. Extensions to these methods herein create signatures based on gene expression, which contain various unique properties and can be correlated to outcomes of interest such as treatment response.

Testing these methods to detect biomarkers of response is challenging due to a lack of real-world data from drug trials. However, a clinical trial was conducted by Pfizer to test avelumab plus axitnib in patients with advanced renal cell carcinoma as part of the JAVELIN Renal 101 program. The Phase III trial reached clinical endpoints and the drug combination was approved. The data from this trial was released publicly, and includes information on patient clinical features and outcomes, as well as high-throughput data such as gene expression, mutation, HLA, etc. The release of this data provides an ideal test case for using geometrical methods to predict treatment response in an oncology clincial trial.

The objectives of this study are to create gene expression signatures, to create and apply a new methodology to the Pfizer clinical trial to determine if subgroups of responders can be identified based these signatures, and to determine if unbiased prediction of samples response to the intervention can be made with high accuracy.

## 2 Materials and methods

### 2.1 Pfizer JAVELIN Renal 101 trial

The Pfizer JAVELIN program consisted of long-term monitoring of patients with advanced renal cell carcinoma [14]. A Phase III trial in avelumab plus axitinib was conducted with a comparator arm of sunitinib as the standard of care. The total number of patients enrolled was 886, with 442 assigned to the treatment arm and 444 assigned to the sunitinib comparator arm.

Data for this trial was provided for multiple data types, including 726 for which gene expression data was available along with clinical parameters and outcomes. Gene expression was given in the form of log_2_ TPM, ie. Transcripts Per Million, with minimum values set to 0.01. Data were not further processed in any way before being used in topological data analysis.

Additional data such as gene mutations, HLA, pathway, etc. were not considered in this analysis.

Clinical information included baseline parameters such as age, PD-L1 positive status (PD-L1+), sex, time to PFS, and binary indication of censor at PFS (that is, inverse of event at PFS). The median time to PFS was reported as 13.3 months in the treatment arm and 8.0 in the comparator arm, resulting in a stratified HR of 0.69 (*p*-value *<* 0.0001). However, it must be noted that in the data provided by Pfizer, using data from the 726 individuals with both clinical and gene expression data, median time to PFS is 4.07 months between arms (8.38 in the comparator arm and 12.45 in the treatment arm) as opposed to the 5.3 months reported. We therefore used the data in its current form, while considering the more modest difference in median time to PFS present for the present study. The KM estimator of the dataset used is presented in 1a.

### 2.2 TDA approach

In this article we build from the local approach as described in [15]. This process assigns to every element in a distance space, a non-increasing function called a stable rank, according to some predefined parameters. A key feature of this signature of a point in a distance space is that it captures some geometrical properties of the embedding of this point into the entire distance space. In particular, points whose embeddings have similar geometrical properties have similar stable ranks, and the embeddings of points whose stable ranks are far apart have different geometrical properties.

The first step of our analysis is to choose a collection of sets of parameters (called geometrical parameters) for extracting stable ranks. There are two kinds of parameters, one related to homology, and one related to the geometry determined by the distance. Geometrical properties are encoded by specifying distributions characterizing various regions of the distance space. For example, for a given point we might explore the geometry of its close neighborhood, or we might explore the geometry of points that are at certain range of distances form the given point.

Extracting stable ranks, with respect to each set of geometrical parameters, is the second step of our analysis. In the third step, for every set of geometrical parameters, we consider the distribution of integrals of the obtained stable ranks. We think about these integrals as a filter function and apply local clustering according to this filter and the *l*_2_ distance in a process similar to the mapper procedure [7]. In this way, for every set of geometrical parameters, we construct various subgroups of the obtained stable ranks, and hence subgroups of the points in the distance space from which we started.

Thus the points in the distance space are grouped according to the geometrical properties of their embeddings as captured by the obtained stable ranks: the embedings of the points in different subgroups have different geometrical character and the embeddings of the points in the same subgroup have similar geometrical character.

We applied this three step process to the gene expression data with the distance given by the Euclidean metric and 14 standard sets of geometrical parameters in each of homological degrees 0 and 1. The outcome is a collection of subgroupings of patients, also referred to as blocks, based on geometrical similarities of the embeddings of the associated gene expression in the Euclidean distance space given by the entire gene expression data. These subgroups are organized into 28 groups, corresponding to our choice of sets of geometrical parameters.

### 2.3 Determination of responders

A key innovation of this paper is a method to determine the classification of individuals, or samples, as a responder or non-responder. The method is applicable to all patients, regardless of censoring or outcome, so that no classifications are chosen based on ie. reaching the event or censoring at an earlier or later time point.

The procedure is conducted in the following iterative manner for each and every sample in the dataset. First, subgroups are formed based on the Euclidean geometry of the expression data across all the sets of chosen geometrical parameters as described in 2.2. Next, for each sample, all the subgroups containing it are combined into a synthetic block by taking the union. The synthetic block’s Kaplan-Meier estimator is then itself tested via log-rank test. The p-value of this test is compared to a threshold, which is based on the p-value of the overall dataset, and can be adjusted as required. In this investigation, a threshold of 0.0001 was employed.

Each datapoint whose synthetic block’s KM is under this threshold is then categorized as a responder, with those failing to reach this threshold categorized as a non-responder. This procedure is conducted for all samples, after which the now classified responders and non-responders can be grouped and tested. Each and every patient in the trial is thus assigned a classification, even if the patient was in the control or comparator arm, or was censored at some timepoint.

This method is not designed to be unbiased, but rather as a method to give “gold standard” classification of responsers and non-responders. In fact, this represents an idealistic partitioning of response classification since it utilizes a p-value threshold of synthetic blocks created via the sample’s membership.

### 2.4 Classification Trees

To determine the weights of features (genes) most strongly represented as predictors of response vs non-response, classification trees were employed. These were built on the entirety of the data, using the classification of responder vs. non-responder as indicated. This was done on all patients to determine which genes might be used as biomarkers of response, using the gold standard of response which may be more stable than other metrics, due to the use of geometry to construct it. In-sample accuracy was determined with the top 10 features, and the top rules for classification were mined.

A prediction of utilizing classification trees on the data was also tested, using the top 10 genes, in order to determine if biomarker partitioning might be a reliable method. This was not testing out of sample, and as such represents internal accuracy and is not unbiased.

Decision trees were created using the python module scikit-learn version 1.7.

### 2.5 Responder testing

To determine the utility of these methods to predict patients according to response, leave one out (LOO) cross-validation was conducted. The purpose was two-fold: first to compare out-of-sample prediction with the response classification, and second to measure the treatment effect captured by unbiased predictions. Each sample was completely removed and the procedure as stated was applied to form subgroups for the rest of the cohort (725 individuals), to create a baseline for comparison. This ensure that the entire process, from measurement of geometry to the formation of the subgroups and of the synthetic blocks, was conducted without the sample test point and thus was strictly unbiased.

The test procedure continued by creating a synthetic block for the testing sample in the following manner. First, using the same procedure as for classification, the geometrical representations of the training data (n=725) was created. Next, the excluded testing sample was transformed into stable ranks. Second, for each set of geometrical parameters one sample from the 725 individuals was chosen whose corresponding stable rank was the nearest (in *l*_2_ distance) to the stable rank of the testing sample. A synthetic block was then formed as the union of the blocks of these nearest chosen samples across all the sets of geometrical parameters. After this procedure was completed, the KM of the synthetic block was tested using the log-rank test, and a similar threshold of p<0.0001 was used to make a prediction of the testing sample as being a responder or not. This was repeated for all samples, giving an out-of-sample prediction for every sample.

An unbiased prediction of out-of-sample accuracy compared to the gold standard classification of subgroups was then possible by comparing the predictions to the previously determined responder classification. Then, the performance of the predictions was measured by creating KM plots of treatment vs comparator arms for each of the responder and non-responder prediction groups. The Restricted Mean Survival Time (RMST) was computed for each curve using the R package survRM2, with a tau of 13 which corresponds to the minimum follow up time specified in the Phase III study, since the largest time at which at least 10% of participants remained at risk in each treatment arm was slightly less. RMST between the two prediction groups was then compared.

It is important to note that the classification methodology and response prediction were designed to be independent procedures. For instance, obtaining unbiased response prediction does not require a classification procedure, nor does measuring concordance with classification labels need to be conducted.

### 2.6 Statistical analysis

The significance threshold alpha was set to 0.05 for all testing of baseline characteristics. The Wilcoxon rank sum test was used for age and Pearson’s Chi-squared test were used where category counts were present.

## 3 Results

The determination of responders resulted in 397 individuals being classifed as responders and 329 classified as non-responders. The KM of this parititioning in Figure 1b and Figure 1c illustrates that it represents a dramatic difference between the groups. Median time to PFS has been radically altered, with the treated groups not reaching the 50% level. Furthermore in the non-responders the avelumab plus axitinib group had markedly worse response than the comparator arm, indicating a directional change.

**Figure 1.**
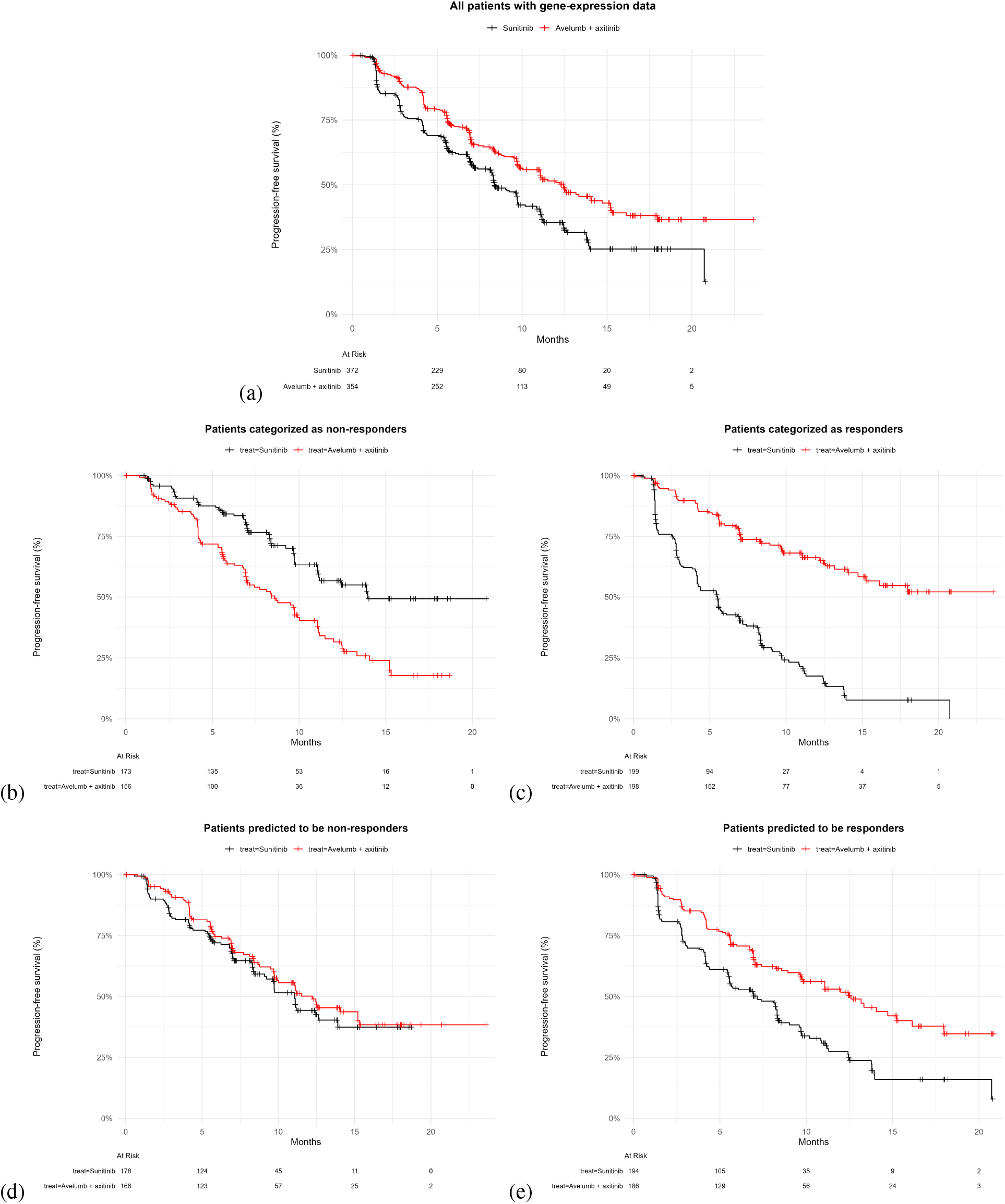
Kaplan-Meier estimator of all patients, by responder categorization, and by responder prediction. (a) All patients in the original study with gene expression data available (n=726) (b) Patients classified as non-responders using geometrical techniques (n=329) (c) Patients classified as responders using geometrical techniques (n=397) (d) Out-of-sample patients predicted to be non-responders using geometrical techniques (n=346) (e) Out-of-sample patient predicted to be responders using geometrical techniques (n=380)

The relative weight of the top genes by importance to paritioning as responder/non-responder is shown in Figure 2. PODXL had nearly triple the value of the next gene, SPRN. Other genes had diminishing weights without a similarly noticable dropoff.

**Figure 2.**
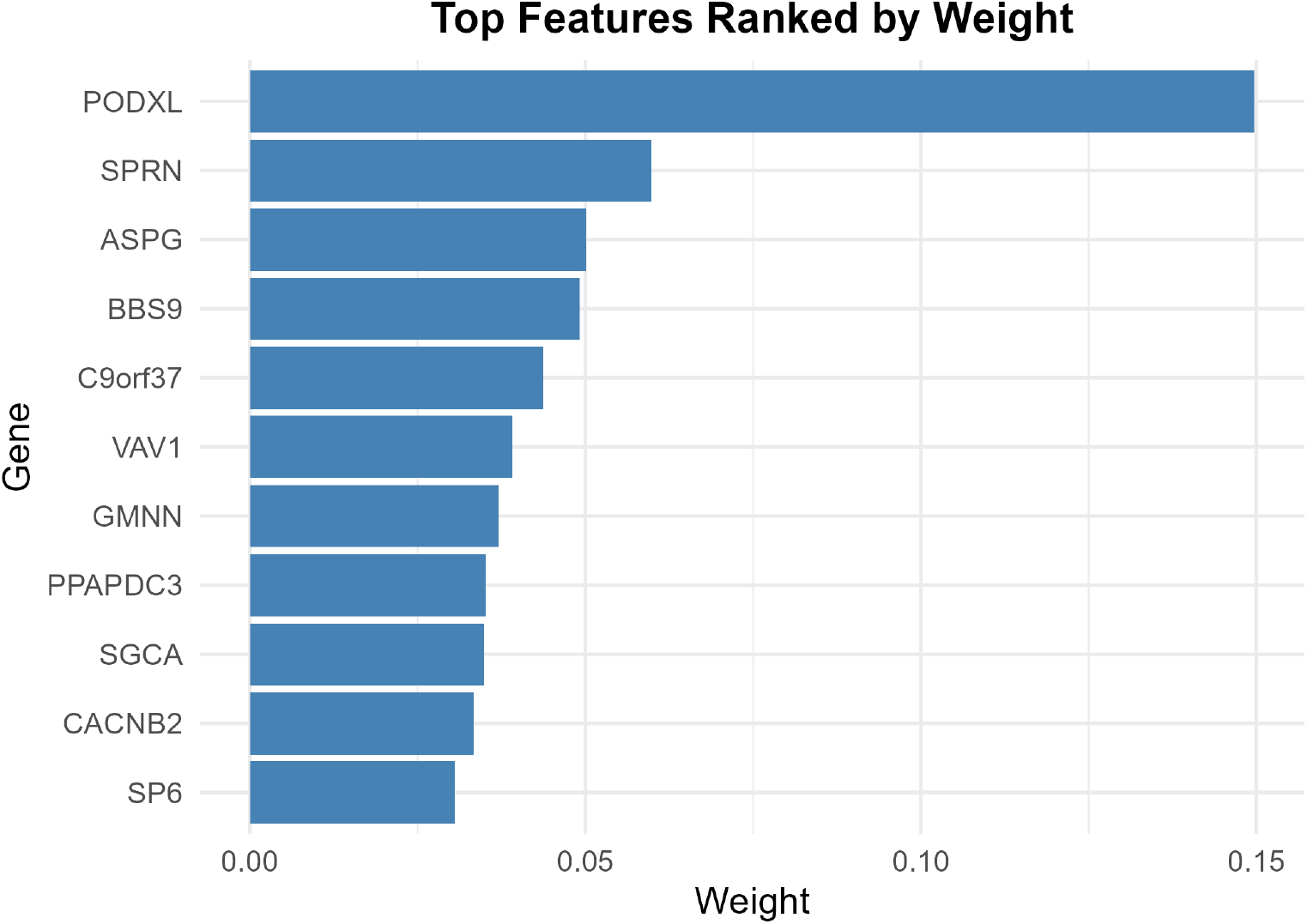
The top gene ranks and relative weights extracted by decision tree classifers on internal response classification.

The classification of responders and clinical characteristics by response group are shown in Table 1. The only characteristic which was significantly different between classified responders and non-responders was age, with responders being 3 years younger than non-responders (p<0.001), which may be a consequence of older patients faring worse in general with cancer and treatment, as well as having higher risk of mortality. Neither gender or PD-L1 status were significantly associated with response classification.

**Table 1:** Baseline characteristcs of the dataset by treatment, and by response classification. The p-values are calculated using Wilcoxon rank sum and Pearson’s Chi-squared tests.

| Characteristic | By Treatment |  |  | By Response |  |  |
| --- | --- | --- | --- | --- | --- | --- |
|  | Sunitinib<br>n = 372 | Avelumab + axitinib<br>n = 354 | p-value | Non-responders<br>n = 329 | Responders<br>n = 397 | p-value |
| **Age** | 61 (53.5, 67) | 62.5 (55, 68) | 0.2 | 63 (56, 69) | 60 (53, 66) | <0.001 |
| **Gender** |  |  | 0.078 |  |  | 0.7 |
| Male | 291 (78.2%) | 257 (72.6%) |  | 246 (74.8%) | 302 (76.1%) |  |
| Female | 81 (21.8%) | 97 (27.4%) |  | 83 (25.2%) | 95 (23.9%) |  |
| **PD-L1 status** |  |  | 0.3 |  |  | 0.11 |
| Negative | 101 (27.2%) | 109 (30.8%) |  | 105 (31.9%) | 105 (26.4%) |  |
| Positive | 271 (72.8%) | 245 (69.2%) |  | 224 (68.1%) | 292 (73.6%) |  |

Prediction of all samples using LOO methodology resulted in 82.0% accuracy. The was consistent among treatment arms. The KM of patients predicted to be non-responders and responders is shown in Figures 1d and 1e, respectively.

The RMST for these groups indicate a mean treatment effect of 2.1 months in predicted responders and 0.55 months in predicted non-responders. In aggregate, predicted responders accounted for 79.2% of the overall treatment effect.

Table 2 illustrates the rules created by pruned trees, with respect to prediction accuracy in-sample, for the responder classification created. Using the top 10 features, 75.9% prediction accuracy was obtained. Since this internal prediction was not designed to be unbiased, it represents the upper limit of prediction possible using this method.

**Table 2:** The top rule sets generated from the classification of responders and non-responders, from in sample construction.

| Pruned Rules for Response Prediction |  |  |
| --- | --- | --- |
| In-sample accuracy with top 10 features: 75.9% |  |  |
| Rule | Conditions | Predicted Class |
| 1 | $PODXL \leq 5.1250$ and $ASPG \leq 4.1100$ | Responder |
| 2 | $PODXL \leq 5.1250$ and $ASPG > 4.1100$ | Non-responder |
| 3 | $PODXL > 5.1250$ and $SPRN \leq 0.9950$ AND $BBS9 \leq 5.5550$ | Responder |
| 4 | $PODXL > 5.1250$ and $SPRN \leq 0.9950$ and $BBS9 > 5.5550$ | Non-responder |
| 5 | $PODXL > 5.1250$ and $SPRN > 0.9950$ and $CACNB2 \leq 2.0150$ | Non-responder |
| 6 | $PODXL > 5.1250$ and $SPRN > 0.9950$ and $CACNB2 > 2.0150$ | Responder |

## 4 Discussion

We have presented a method for representing data points through signatures encoding geometrical properties of how datapoints are embedded in a data set. This has allowed us to build machinery for comparison of gene-expression signatures within a clinical trial to identify the “gold standard” responders as well as predict individual response with high accuracy. Our predictions stratify responders with nearly 80% of the treatment effect, which as noted is conducted in an unbiased out-of-sample manner not reliant on the response classification. This use case shows strong utility of our method for cancer trial data and precision oncology, ie. getting the correct treatment to the correct patient based on individual measurements.

The methods we have developed first transform the data and then analyze the obtained signatures. These signatures integrate certain geometrical aspects of how individual points embed into the space of gene expression data. This has key advantages over standard machine learning (ML) and (artificial intelligence) AI implementations applied directly to the data. For example, our method is able to find results with only a few hundred data points when the number of features is on the order of hundreds of thousands. Typically, ML and AI techniques requires a large number of datapoints to train models, and the presence of huge numbers of variables creates a noisy point cloud where extraneous features may reduce model performance. This is in contrast to our TDA method which has denoising properties, enabling to identify signals of otherwise non-observable geometrical patterns.

The geometrical representation of the data according the the method utilized here inherently anonymizes data. For example, the geometrically transformed gene expression signatures cannot be reconverted to raw data by any means. This transformation can therefore be used without revealing patient data if it is sensitive in nature. While largely not a concern with tumor gene expression, more sensitive data such as genomics may theoretically be used to re-identify individuals. This is also a useful additional component if data must be transferred across various country of origin lines, where legal regulations may forbid the use of identifiable data, or when the data contains intellectual property to remain private.

Current methods in precision medicine for cancer typically prioritize mutations and addressable oncogenes, which derives from basic research and a need to identify the biological mechanisms underlying model of action of a compound. However, as the dependency map project illustrated, gene expression shows the strongest correlation with treatment response in cell lines [6]. Therefore, we take a more holistic view of determining treatment response from the aggregation of all gene expression data without consideration of underlying biological mechanisms. The use of the measurements at all genes leverages the power of geometry where complicated relationships between variables may exist and are difficult to model, and where the use of some subset of variables is likely to reduce signal substantially. Because our geometrical signatures are built in a completely unsupervised fashion and without any input from treatment response, these transformations are robust and downstream correlation with treatment is therefore unbiased.

There are three clear benefits to utilizing this approach. First, it allows for classification of all patients into responders and non-responders, regardless of the intervention given or the outcome with respect to the event or censoring. In the original study, only 157 patients (21.6%) were treated and reached the endpoint of PFS. These might be stratified based on median outcome, although most differential analyses tends to only use quartiles. An additional 85 patients (11.7%) were censored after the midpoint in time between KM curves (10.42 months), and might be reasonably classified as responders. Our method allows for full classification and removes any need for delicacy when a patient outcome or censoring might leave the response level in question. Second, the creation of biomarkers based on this approach are far more likely to be stable than doing a differential gene (DE) analysis. With over 200 thousand genes routinely measured in RNA-seq, the variability in measurement tends to fill the expected distribution, so that any associations are likely to be spurious. In a DE analysis, the gene level difference is the justification to select the marker, while geometrical analysis provides a reference to the relationship between samples before then determining specific markers that might have stable correlation with the groupings determined by the geometry. This increases the likelihood that a biomarker finding may be prognostic in nature. Third, as the high prediction accuracy demonstrates, the geometry is associated to the outcome of interest, namely drug response. This also reinforces that the biomarkers detected are a result of the geometry, and thus related to response instead of simply being randomly distributed.

Another key finding of our investigation is the decision tree of genes contributing to response in the classified groups. The strongest predictive gene, PODXL (podocalyxin), is expressed in multiple cell types including in kidney podocytes [16]. It has been shown to be overexpressed in 9.6% of renal cell carcinomas [17]. Several studies have implicated it as a potential prognostic marker of outcomes in various cancers, including renal carcinomas [18]. Interestingly, the authors of that study report that in kidney renal papillary cell carcinoma overexpresion of PODXL was associated with poor survival, while in kidney renal clear cell carcinoma lower expression of PODXL was associated with poor outcomes. This inconsistency was reflected in the resultant rules for the responder subgroups classified, with rules denoting responders with PODXL both over the threshold value of 5.235, and under the same threshold in combination with other gene rules. Simiarly, subgroups of non-responder were also created with PODXL both above and under the generated threshold value. The other genes with lower relative importance do not have strong literature reporting possible oncological prognostics. The ability to generate such gene prioritization lists represents an enormous opportunity to both identify biomarkers of response and also to delve further into the biological mechanisms of drug action.

This study is limited by the accuracy of the data provided by Pfizer in the publication. Any errors in the collection, processing, measurement and analysis of the patient and tumor derived data may reduce the accuracy of our results. Characterization of clinical variables, such as the primary outcome PFS or times to censor may likewise alter results. Given that this is a published Phase III clinical trial, it is assumed that these errors will be minimized to those that are present in any strictly controlled study.

These methods can be applied in Phase II trials to determine inclusion/exclusion criteria for Phase III studies, as well as to create additional primary or secondary endpoints for analysis. This might increase the chances of passing Phase III substantially. In failed Phase III trials, these techniques may be able to detect similar subgroups of responders not visible with the analytical methods used at the time of the trial. This represents an enormous opportunity, since thousands of oncology drugs have failed Phase III trials, the majority due to lack of efficacy. Identifying the most efficacious subgroups in a post-mortem fashion may allow for application of a fast-tracked trial to confirm the finding. This could potentially bring a large number of previous failed cancer drugs to market as well as improving the treatment effect via subgroups of response, thereby benefiting patients suffering from an incurable disease. And as previously suggested, this method or biomarkers from these analyses could be used to create companion diagnostics in order to predict which patients might have the best response to therapy in approved drugs.

Repeating this procedure on additional Phase III data is desirable, but difficult due to a lack of publicly available datasets. Initiation of collaborations with pharmaceutical consortia may be effective at creating a larger scale testing framework. Such efforts may be utilized not only in single compounds, but also across multiple oncology clinical trials to identify expanded use cases and potential drug repurposing across oncology phenotypes.

## 5 Conclusion

The application of our gene expression signatures and geometrical methods on Pfizer avelumab plus axitinib data identified both gold standard classification of responders as well as unbiased out-of-sample prediction accuracy of 82.0% to identify responders. This accounted for 79.2% of the treatment effect, thus indicating that the methodology has clear utility in oncology clinical trials.

While this clinical trial met primary endpoints, in Phase II and Phase III studies the ability to both identify and detect subgroups of response may dramatically improve chances of a compound passing clinical trials, and these methods may be used as a companion diagnostic or to identify biomarkers of response.

## Data Availability

The data in the present study were open-sourced by Pfizer and available online at: https://www.ncbi.nlm.nih.gov/pmc/articles/PMC8493486/

## 6 Data and software availability

Pfizer’s avlemuab plus axitinib clinical trial data are freely available at: https://www.ncbi.nlm.nih.gov/pmc/articles/PMC8493486/ under “Supplemental Materials”. The analyses conducted in this paper are part of a proprietary codebase which is closed source and therefore not publicly available. A web application which will allow for some of the analyses contained within is under preparation.

## 7 Acknowledgements

WC was partially supported by the Wallenberg AI, Autonomous System and Software Program (WASP) funded by Knut and Alice Wallenberg Foundation, and MultipleMS funded by the European Union under the Horizon 2020 program, grant agreement 733,161. Both WC and RR own equity in The Responder Lab (DatAnon Corporation), which owns the codebase on which this platform has been created as well as related intellectual property. WC and RR have applied for patents on some of these methods.

## 8 Author contribution

WC and RR designed the study, conducted experiments, analyzed results, and wrote the paper.

